# Protocol and rationale for the International PANS Registry (IPR; pediatric acute onset neuropsychiatric syndrome)

**DOI:** 10.1101/2023.09.15.23295605

**Authors:** Erin E. Masterson, Jessica M. Gavin

## Abstract

The International PANS Registry (IPR) is the first centralized, epidemiologic database of children with Pediatric Acute-Onset Neuropsychiatric Syndrome (PANS) and PANS-like features and their siblings. PANS is a relatively new umbrella syndrome that lacks diagnostic biomarkers and is characterized by a set of working criteria. The failure to find a diagnostic biomarker is likely due to underpowered studies and inherent biological heterogeneity within PANS. Until the IPR was established, a critical barrier to large-scale longitudinal studies had been the absence of a large-scale epidemiologic study and a centralized database of children with PANS and PANS-like features. The IPR was created to serve as a translational health tool to accelerate research on the broad spectrum of complex pediatric neuroimmune conditions with the long-term goal of enabling a paradigm shift in this field from symptom-based evaluation and treatment towards biology-based diagnoses, treatments, screening, and surveillance. To date, the IPR has registered 1,666 families (3,247 children) and is the largest database in the world that gathers in-depth information on children with PANS and PANS-like features and their siblings. Enrollment in the IPR is open and ongoing; longitudinal follow up is planned. Participating families enroll their children with PANS and PANS-like features and their healthy siblings in the IPR via an online survey platform. The selection criteria for IPR enrollment are intentionally less restrictive than the current working criteria for PANS to generate a large recruitment pool and enable study of the broad spectrum of PANS-like conditions. The IPR is designed to enable ancillary study recruitment based on detailed selection criteria and to grow and expand in scope in the future. The IPR team is committed to data sharing and invites collaborators who will leverage existing data from the IPR database and extend knowledge in an area beyond the original scope of the IPR.

## Introduction

Pediatric Acute-onset Neuropsychiatric Syndrome (PANS) is relatively newly defined and is a diagnosis of exclusion, characterized by acute onset of Obsessive-Compulsive Disorder (OCD) and/or severely-restricted food intake and at least two other neuropsychiatric symptoms in previously healthy children (1–4). These may include anxiety, emotional lability, irritability/aggression, behavioral regression, deterioration in school performance, sensory and/or motor abnormalities, and somatic signs and symptoms (1). Importantly, PANS does not have a validated diagnostic biomarker or ICD-11 diagnostic code (5). PANS is currently diagnosed using non-validated working criteria, putting patients at risk of misdiagnosis, delayed treatment, or inappropriate therapeutic intervention. In some cases, long-term psychiatric and neurological impairment; academic, developmental and behavioral regression; and severe social and emotional disruption to patients and their families may occur (4,6–10). The failure to find a diagnostic biomarker is likely due to underpowered studies and inherent biological heterogeneity within PANS.

A leading hypothesis is that the illness is driven by a brain-reactive autoimmune or neuroinflammatory response to infection (4,6,11). PANS is likely an umbrella syndrome comprised of subgroups (7,12), the most well-known of which is Pediatric Autoimmune Neuropsychiatric Disorders Associated with Streptococcal Infections (PANDAS) (13–15). PANS and PANDAS patients tend to experience relapsing episodes of severe neuropsychiatric symptoms (16), but prospective studies testing for temporal correlations between symptoms and infection have yielded conflicting results—likely owing to small sample size (17,18). While some larger observational case-control studies support an association between infection and PANDAS (19), intra-individual analyses have not been possible. PANS has been observed in siblings and family clusters, suggesting genetic and/or environmental subtypes of PANS (20–23). Indeed, rare variant and genotype studies support a genetic basis for at least some subgroups of PANS (24,25). Furthermore, it is possible that PANS etiology may be connected to the prenatal environment, related to maternal immune health (25–28).

Until recently, a critical barrier to large-scale longitudinal studies had been the absence of a standardized, central database of PANS patients. The University of Washington’s (UW) International PANS Registry, launched on July 19, 2020, has registered 1,666 families (3,247 children) to date and is the largest database in the world that gathers in-depth information on children with PANS and PANS-like features and their siblings.(29) The IPR fulfills a long-standing call in the clinical and scientific communities for large-scale epidemiologic study and a centralized registry for children with PANS and PANS-like features (4). The IPR was created to serve as a centralized hub of information and translational health tool to accelerate research on the broad spectrum of PANS-like conditions with the long-term goal of enabling a paradigm shift in this field from symptom-based evaluation and treatment towards biology-based diagnoses, treatments, screening, and surveillance. The IPR was designed to expand globally and grow so as to represent a large and diverse participant pool, to enable longitudinal studies, and to serve as a recruitment tool for clinical trials and subsequent, ancillary studies—particularly those focused on more homogenous subgroups. Ultimately, the IPR aims to improve diagnosis and accelerate therapeutic discovery by facilitating large-scale biomarker discovery and mechanistic research of this complex illness, including identifying underlying subgroups and their dimensions.

## Methods and analysis

### Study design

The International PANS Registry (IPR; www.pansregistry.org)(29) is an open patient registry housed at the University of Washington (UW), established in 2020 and sponsored by the Pediatric Research & Advocacy Initiative (PRAI), a 501(c)3 parent-led organization (29,30). The IPR is the first large-scale, epidemiological database in the world for PANS and related neuroimmune conditions. The IPR was also designed to be able to expand globally and grow so as to represent a diverse and sizeable participant pool, to enable longitudinal follow up, and to serve as a recruitment tool for subsequent studies. All participants have consented to be followed longitudinally and to receive invitations to participate in subsequent and/or ancillary studies. To date, the IPR data is observational, survey data based on parent report and includes cases and healthy siblings, who we do *not* consider as study controls. Future goals include linking existing data to ancillary studies and expanding the scope of included data elements by enrolling study controls, integrating pointed medical record information, and creating a linked biobank to enable the evaluation of potential disease biomarkers and genetic information.

### Patient and public involvement

The IPR is a collaborative community-based research partnership between the two authors, an epidemiologist (EEM) and the director of the PRAI (JG), the IPR sponsor. PRAI, a nonprofit organization supported by the parent/patient community raised funds from families affected by PANS and related conditions to fund the creation of the IPR, the first registry for families affected by PANS and related conditions. Through social media platforms, thousands of families affected by PANS from around the world contributed to the development of the IPR survey content. These efforts reflect the patient community’s commitment to the project and their confidence that the IPR could accelerate research and elucidate the barriers to children receiving timely and appropriate diagnoses and treatment.

### Participants

The target population for the IPR is children, adolescents, and adults in the U.S. and Canada with PANS-like neuroimmune conditions (cases) and their siblings who were not believed to be affected by these conditions at the time of enrollment. Enrollment in the IPR is ongoing and open to the general public via the IPR’s website (www.pansregistry.org). Adult caregivers enroll and respond to all survey questions for their child(ren) less than 18 years of age or who are adults for whom the caregiver has a formal Power of Attorney.

#### Eligibility criteria

We invited families who believe their previously healthy children, who did not have chronic neurological and psychiatric problems, developed PANS-like features they believe are related to an immune response, irrespective of a PANS diagnosis by a clinician. Individuals in recovery as well as those with active symptoms at the time of enrollment are eligible. We did not restrict IPR enrollment eligibility to the current PANS working criteria (1,4). Given that PANDAS is considered a subgroup of PANS (4), individuals suspected of having PANDAS are included in the study population. We intentionally cast a wide net for inclusion to capture those with milder symptoms and insidious onsets to generate a large recruitment pool by which immune-mediated neuropsychiatric health may be broadly studied. To this end, we also included siblings who have not experienced any PANS-related early signs or symptoms based on the knowledge that cases cluster in families/ genetic component.

To focus on those with conditions that lack a diagnostic biomarker and are not classified as a known illness or disease, participants who had received an autoimmune encephalitis (AE) diagnosis *before* their reported onset of PANS symptoms were not eligible to participate during the first enrollment phase (July 19, 2020 through December 2021); this exclusion criteria was dropped for recruitment beginning on July 19, 2022 to accommodate the possibility that PANS has been misdiagnosed as AE and to ensure we’ve included these individuals. We asked detailed questions about lab results and the basis for diagnosis among those who report having AE so that discretion may be used on a case-by-case basis regarding their inclusion in research studies based on the IPR database.

#### Recruitment strategies

We recruited patients by convenient sampling directing interested parties to the publicly accessible link on the IPR’s website, using Facebook, mailing lists (including emails and newsletters), partnerships with other non-profit organizations, clinical referral networks, local community events, and international conferences. JG led these recruitment efforts.

### Survey Development

The IPR surveys originated from engagement between clinicians, researchers, and more than 1,500 members of the online PANS parent community, led by JG. Some of the survey questions emerged from social media interactions and informal polling in online parent support groups, which are platforms that have allowed thousands of families from around the world to compare notes on common clinical reports, histories, symptom presentations, and treatment responses they have observed in their children. EEM, an epidemiologist, worked with JG to organize and convert these topics and questions into the set of surveys listed in Table 1, which comprise the baseline IPR data. During early 2020, we conducted an iterative piloting process of the full set of surveys with the families of 100 volunteer cases and their siblings.

**Table 1.**
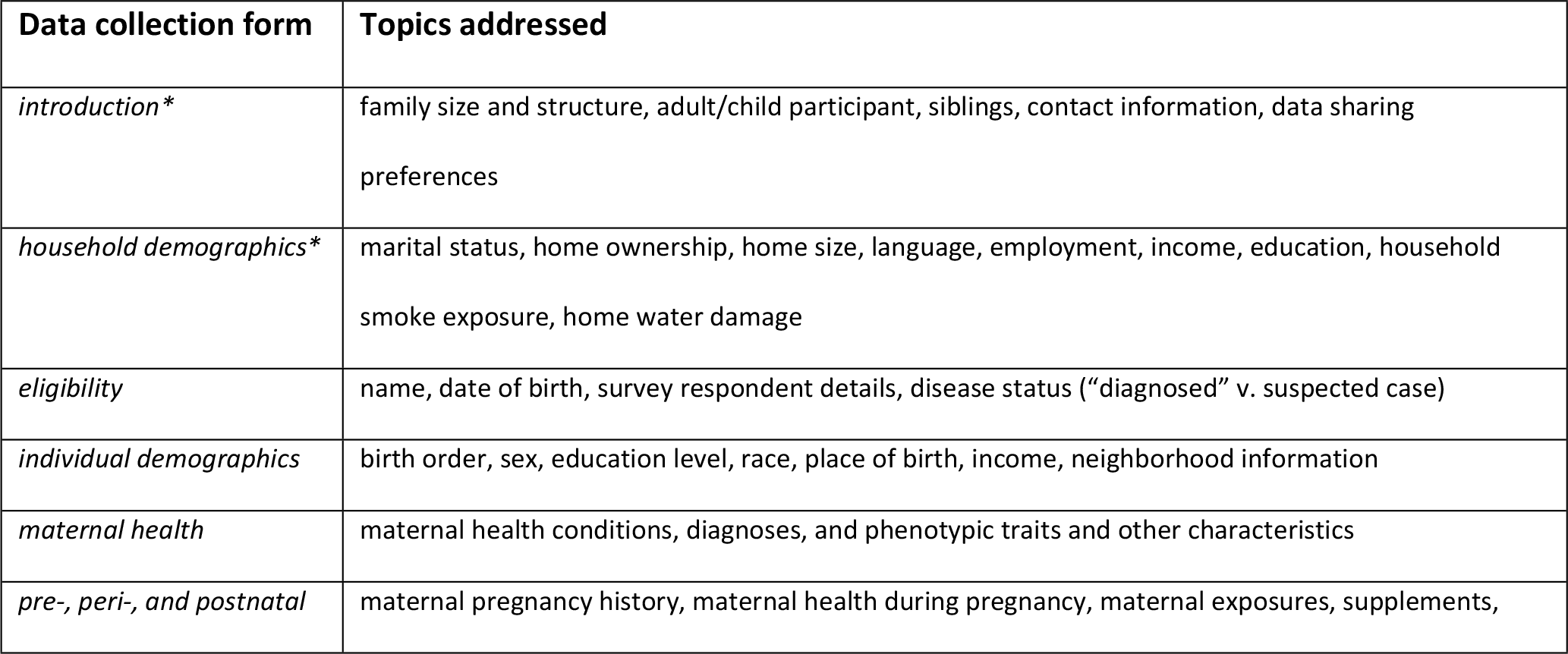
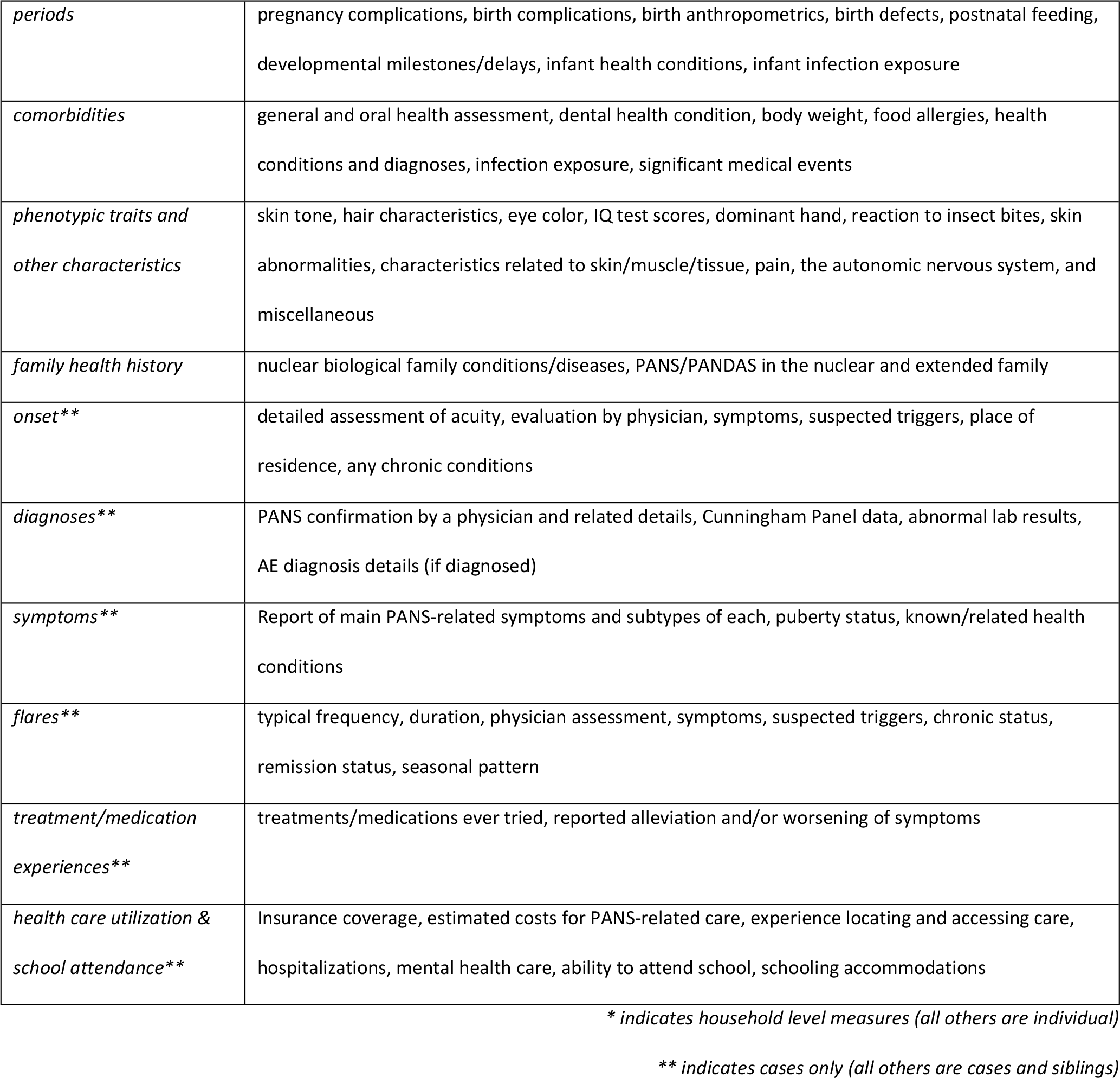
Overview of baseline survey forms for the International PANS Registry.

### Data elements

IPR’s baseline survey questions address the following topics: eligibility, demographics, maternal & child health (MCH), patient’s current health status, comorbidities, and characteristics, and family health history. We collect additional information on PANS/PANDAS cases, including details regarding onset, diagnoses, symptoms, flares in symptoms, treatment experience, health care utilization, and school attendance (Table 1). We inquired about symptoms, family health history, infectious disease exposure and immune dysregulation, and general medical history, guided by existing PANS literature and patient advocate websites (1,4,31–33). We also designed our survey about treatment and medication experience to align with clinical management approaches described by PANS experts in the literature.(34– 36) For questions that related to those asked in national surveys, including from the National Health and Nutrition Examination Survey (NHANES) and the Pregnancy Risk Assessment Monitoring System (PRAMS) (37,38), we used the same questions and response options as the national surveys to be able to make comparisons to national population averages. To our knowledge, this study includes detailed data on certain topics that have not been formally collected within this patient population. These questions originated from polls and discussions on patient support group social media pages. The topics relate to phenotypic characteristics (maternal and child), maternal health, early life of the child (pre-, peri-, and postnatal), and early disease experience.

We ask parents to have their child’s medical records, including their vaccine record, clinical test results and lab reports, in hand so that they can accurately report these results in the survey forms. We also capture parent response for each of the current working criteria for PANS and PANDAS (4). The complete set of the first version of surveys constitutes more than 2,000 data fields for cases and more than 1,400 data fields for siblings. Complete data dictionaries describing the baseline survey data available may be found on the IPR website (https://pansregistry.org/researchers/).

### Data collection and storage

IPR data collection began on July 19, 2020 on the Dacima Software Inc© platform. Data collection on this platform concluded in December 2021. In July 2022, IPR data collection and management activities transitioned to the UW and resumed on the Research Electronic Data Capture (REDCap) software platform supported by the UW’s Institute of Translational Health Sciences (ITHS) (39). REDCap (Research Electronic Data Capture) is a secure, web-based application designed to support data capture for research studies, providing: 1) an intuitive interface for validated data entry; 2) audit trails for tracking data manipulation and export procedures; 3) automated export procedures for seamless data downloads to common statistical package; and 4) procedures for importing data from external sources.

The full set of original surveys (on the Dacima platform in 2020-2021) took 2-3 hours per case to complete. In 2022, the baseline surveys were pared down when the survey platform transitioned to REDCap at the UW (40). Completing the full set of this second version of baseline surveys takes less than one hour per case. On both platforms, participants were allowed to save their work and continue in separate segments of time.

Until 2021, any participant who signed up (with their email contact) and provided (a) either name or date of birth and (b) case/sibling status was included in the IPR Recruitment Pool. Beginning in 2022, participants may opt to sign up for recruitment purposes *only* and are asked to complete a 15-minute survey per individual enrollee (includes variables listed in Table 2). For those interested in more in-depth participation, embedded within the IPR is the IPR Epidemiology Study, an observational, longitudinal study that will investigate the epidemiology of the broad spectrum of PANS through a series of analyses and manuscripts led by IPR investigators (manuscript describing baseline characteristics under review).

**Table 2.**
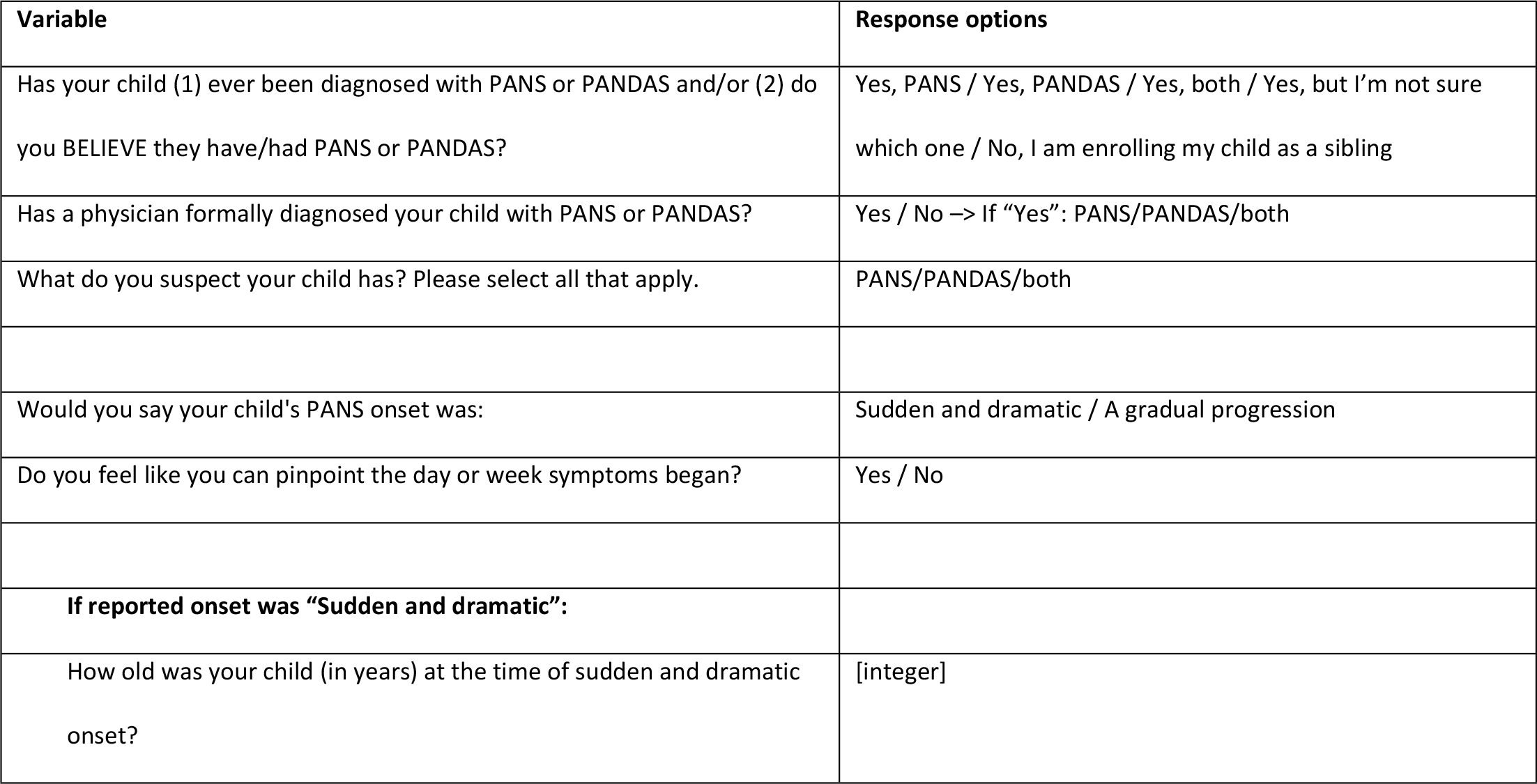
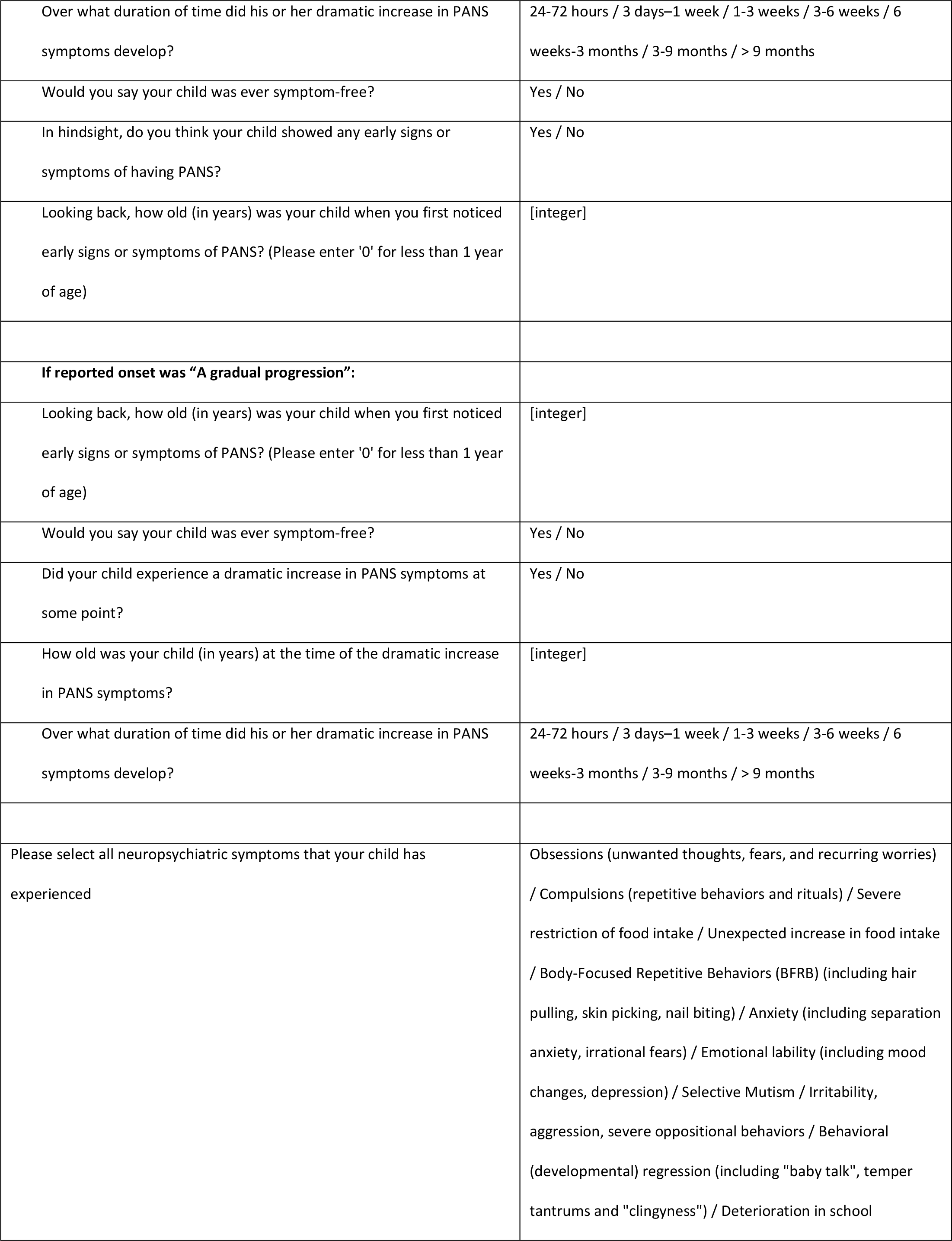
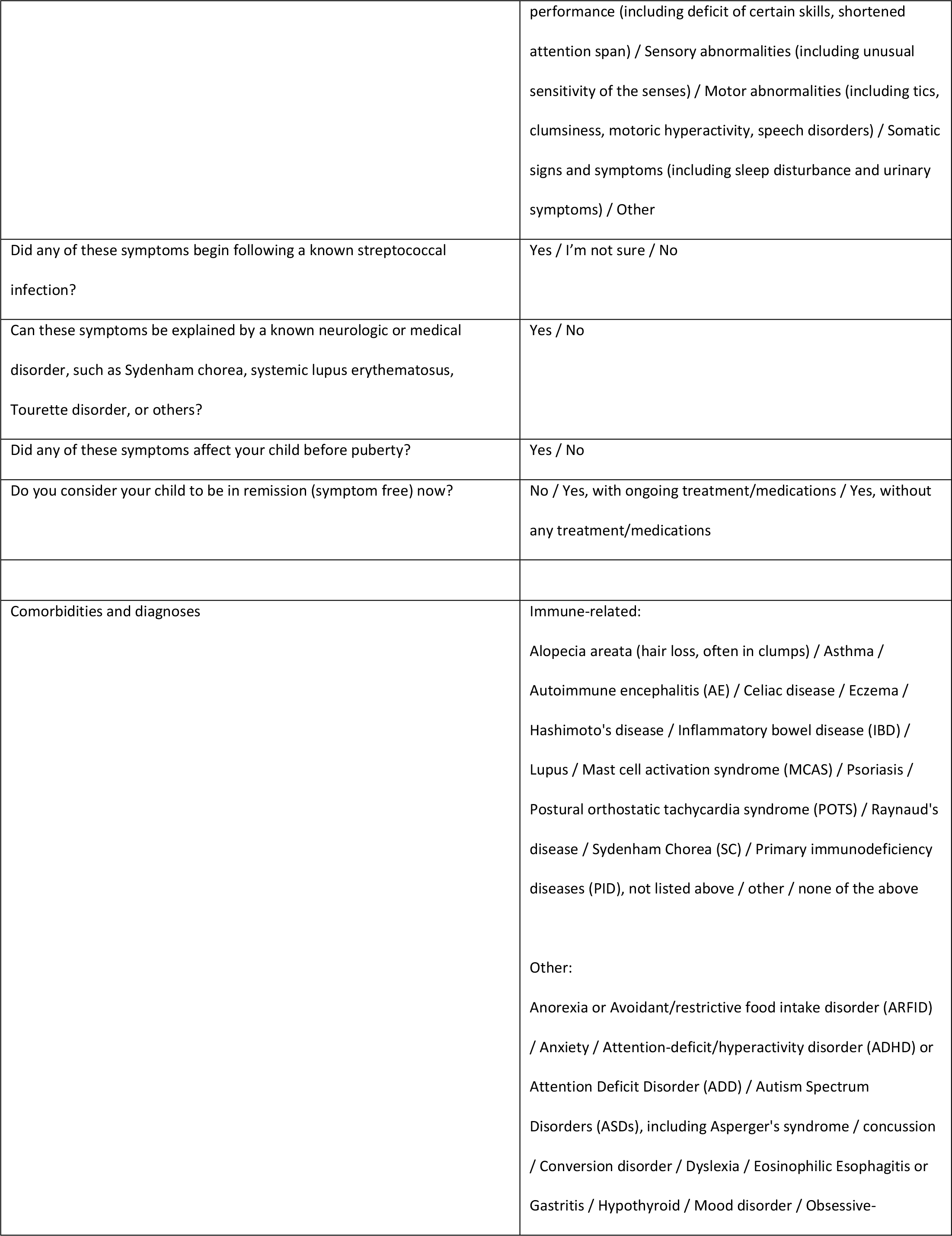
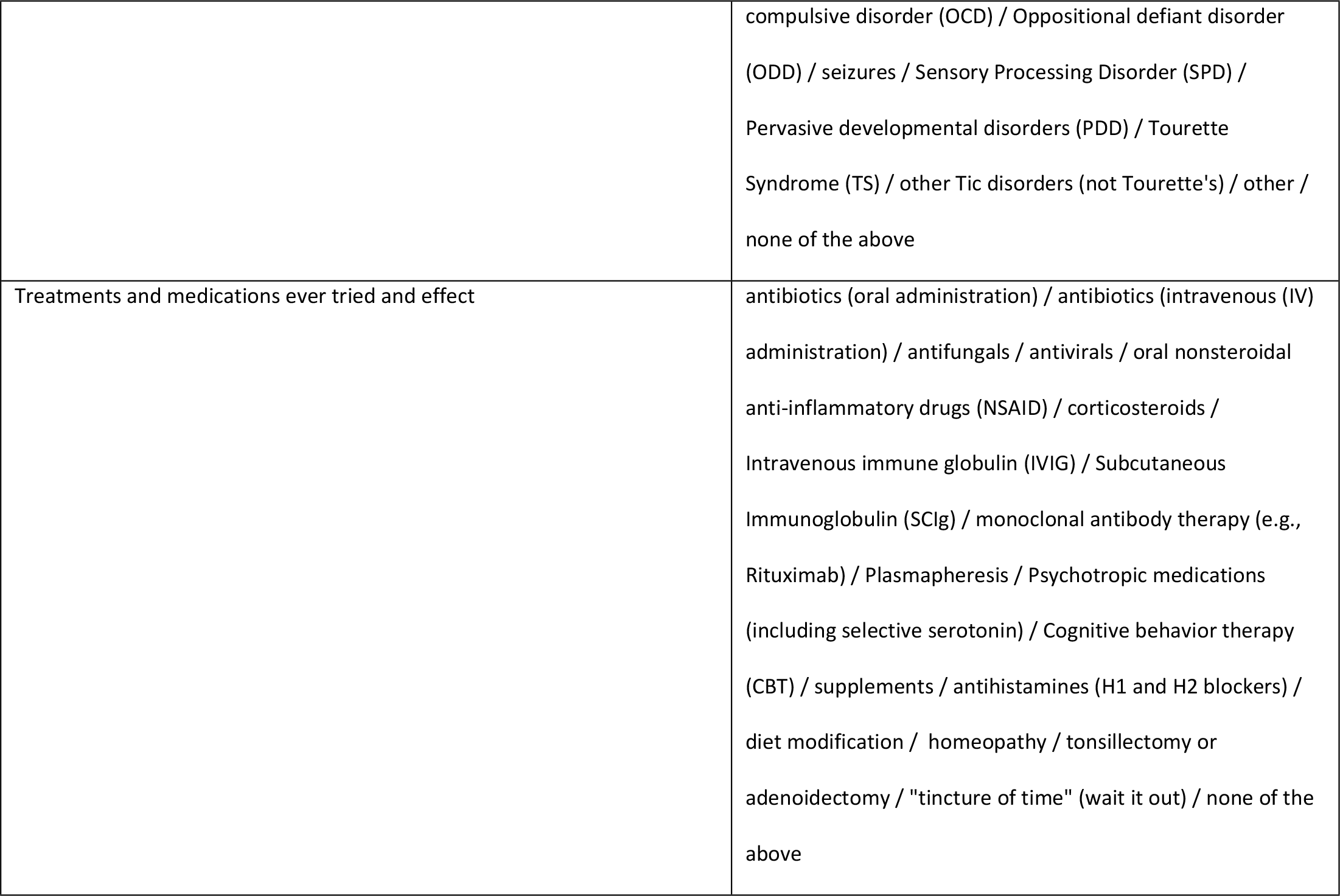
Parent-reported variables available for ancillary study sample selection from the International PANS Registry.

### Analytic plans

IPR survey data will be used to describe clinical, phenotypic, and epidemiologic characteristics of participants and to explore patterns in these characteristics across the IPR sample. The IPR team will analyze baseline survey data using SAS version 9.4 (SAS Institute, Cary, NC, USA). We will clean the data and present descriptive statistics alongside detailed data documentation. In subsequent analyses, we will rely on regression models to evaluate relationships within the data. Analytic techniques to classify subgroups or identify clusters in the data will include latent class analysis and supervised and unsupervised machine learning approaches. We will apply appropriate evaluation metrics depending on the model type. We will consider p-values <0.05 and/or 95% confidence intervals not including 0 value as “statistically significant” and will correct for multiple testing when appropriate.

### Ancillary study subsample selection

Collaborators may target a subsample of the IPR for ancillary study recruitment by applying various criteria to identify narrow, homogenous, and moderate/severe participant subgroups. Table 2 lists survey questions that describe participant diagnoses, symptom onset characteristics, and treatment/medication attempts. While we expect these to be the most commonly relied upon selection criteria, all variables listed in Table 1 above may also be utilized for sample selection purposes.

### Ethics and dissemination

Data collection activities in 2020 and 2021 were approved by the IntegReview (now Avarra) Institutional Review Board (IRB) (#PRAI_svy001) for U.S. and Canadian citizens. Beginning in 2022, data collection activities were additionally approved by the University of Washington (UW) IRB (#STUDY00014294) for U.S. citizens. All participants have consented via an e-signature on survey software platforms to be followed longitudinally and indefinitely, to receive invitations to participate in ancillary studies, and to share their de-identified data. De-identified data from the IPR are available to the public via an online data request portal (https://pansregistry.org/researchers/).

### Status of the IPR recruitment pool

At the end of 2021, the IPR had registered 3,247 children from 1,666 households. Enrollment remains open. These participants included cases and their healthy siblings. Most of the 1,666 households enrolled reported having one to three children (94%). To date, participating households are from all regions of the U.S. and Canada but are not evenly distributed.

## Discussion

To date, the IPR has registered 1,666 families (3,247 children) and is the largest database in the world that gathers in-depth information on children with PANS and PANS-like features and their siblings. This database will serve as a translational health tool for conducting studies that address unresolved research questions around immune-mediated neuropsychiatric conditions related to PANS. The selection criteria for IPR enrollment are intentionally less restrictive than the current working criteria for PANS to generate a large recruitment pool and enable study of the broad spectrum of PANS-like conditions using big data analytics and tracking patients over time. Current limitations of the IPR include that the survey data is based on parental self-report and subject to recall bias. The convenient sample of participants is not generalizable or representative of any region, demographic, nor specific patient population.

In the long term, the IPR hopes to contribute to improving patient care, public health, and policy related to PANS by providing a tool for developing biology-based diagnoses, treatments, screening and surveillance. The IPR maintains open enrollment and is designed to enable ancillary study recruitment based on detailed selection criteria and to grow and expand in scope in the future (e.g., enroll study controls, integrate medical record information, collect biospecimens). To this end, the IPR team is also committed to data sharing. De-identified data from the IPR are available to the public via an online data request portal (https://pansregistry.org/researchers/). Complete data dictionaries describing the survey data available may also be found at this site. Participant data is kept confidential. The data is coded with a non-identifying subject ID. The Registry is intended to serve as a recruitment tool for new studies. We invite researchers to design studies that leverage existing data from the IPR database and extend knowledge in an area beyond the original scope of the IPR, including but not limited to immunology, rheumatology, microbiology, psychiatry, neurology, and genetics.

## Author Contributions

EEM and JG jointly conceptualized the IPR. EEM acquired ethics approvals and led the survey design and writing of the original draft of this manuscript. JG contributed to designing the surveys and was responsible for acquisition of funding, study participant recruitment, and contributed to reviewing and editing this manuscript. Both authors have read and approve of this final version of the manuscript for publication.

## Acknowledgements

The authors thank the participating families for their dedicated time and commitment to their community goal of building the IPR. We appreciate the input of those who helped inform our survey design, the time of those who shared their children’s health information, and the many contributions of those who supported the fundraising effort to support this project. We specifically thank the Neuroimmune Foundation for their key contributions to our recruitment efforts. We would like to acknowledge the support of Dacima Software Inc©, the survey platform on which the first wave of data reported here was collected.

## Ethics approval

Data collection activities in 2020 and 2021 were approved by the IntegReview (now Avarra) Institutional Review Board (IRB) (#PRAI_svy001) for U.S. and Canadian citizens. Starting in 2022, data collection activities were approved by the UW IRB (#STUDY00014294) for U.S. citizens.

## Data availability statement

The IPR team is committed to data sharing. De-identified data from the IPR are available to the public via an online data request portal and subject to an approval process (https://pansregistry.org/researchers/). Participant data is kept confidential. The data is coded with a non-identifying subject ID. Complete data dictionaries describing the survey data available may also be found at this site. The Registry is also intended to serve as a recruitment tool for new studies. Please contact the study PI and lead author of this manuscript to further discuss interest.

Informed consent documentation may be made available upon request. Participants who provided their email address upon enrollment consented to be contacted in the future with messages/updates and subsequent data collection requests from the IPR and, potentially, other researchers who would like to select a study sample from the IPR recruitment pool. The data they provided when they enrolled in the IPR may be used for future studies and recruitment purposes (i.e., study inclusion criteria). We allow participants to select specific permissions for the type of third party with whom they wish their data be shared with when they enroll and will select data accordingly for outside requests.

## Collaborators

The IPR team invites external collaborative research proposals to use the IPR database for analyses and recruitment.

